# A review of scientific theories used to justify the delivery of school-based mindfulness programmes. A protocol for a scoping review

**DOI:** 10.1101/2023.06.19.23291616

**Authors:** Pamela Abbott, Graeme Nixon, Isabel Stanley, Lucia D’Ambruoso

## Abstract

**Background:** Mindfulness-based interventions are increasingly delivered in schools with the aim of promoting the mental wellbeing of pupils. The apparent success of such interventions has led to calls for mindfulness to be included in the school curriculum. However, the findings are more mixed than is often acknowledged and this may in part be because the theories justifying are rarely considered. Understanding the scientific theories underpinning SBMIs may improve the outcomes from interventions. In this paper we outline our scoping review protocol to systematically review the literature to map the scientific theories that have been identified to justify using school-based mindfulness interventions to promote pupils’ wellbeing.

**Methods:** The scoping review methodology will guide the conduct of the review. We will limit our search to scholarly databases and PhD theses to ensure the credibility of the theories. The following databases will be used: MEDLINE, PsychINFO, Web of Science Core Collection, PubMed Central, Scopus and ProQuest Dissertations and Theses. Two reviewers will independently screen all abstracts and full-text studies for inclusion. We will include any study that includes a theoretical explanation linking a mindfulness intervention with its impact. Information from the papers will be extracted using a framework developed by the team.

**Discussion:** The scoping review will map, classify and compare scientific theories that have been used in the literature to justify the use of school-based mindfulness intervention to promote pupils’ mental wellbeing. The findings will be disseminated through a peer -reviewed journal article, conference presentations, our project website and social media.

## Background

The proposed literature review aims to investigate the theoretical frameworks underpinning school-based mindfulness interventions (SBMIs) designed to promote pupils’ mental wellbeing. It is often claimed that mindfulness interventions are informed by theories and practices from contemplative traditions, science, medicine, psychology and education [1–3]. However, the theories underlying them are rarely discussed in the literature, which has focused on determining if they work, that is, if interventions have the intended outcomes. The apparent success of SBMIs in promoting the wellbeing of children and young people, as evidenced by systematic reviews and meta-analyses [4–10], especially when integrated into a whole-school approach [4,7,11], has led to calls for mindfulness to be included in the school curriculum [12–14]. Mindfulness interventions have also been shown to reduce teachers’ stress levels, improve their mental wellbeing and life satisfaction, and their relationships with students making classroom environments more conducive to student learning [13–18].

However, the reviews have been more mixed than is often acknowledged, and claims for the effectiveness of mindfulness may have been exaggerated [19]. The mixed findings from reviews may, at least in part, be due to the heterogeneity in SBMIs in educational settings. There is a lack of agreement on what mindfulness is [1], and programmes differ in content, how they are delivered, who delivers the training and the length and intensity of the training [20]. They can be whole school interventions with mindfulness built into the school curriculum, or sometimes only taught to some years or classes or only used with some children, usually those with behavioural or mental health problems [7,20].

It has been argued that complex interventions based on theory can be more effective [24], enabling an understanding of how the intervention worked, not just if it worked. When a theory of what mindfulness *is* and *does* underlies the intervention, it provides additional insight into the characteristics of effective interventions

The findings will enable us to refine the programme theory for research we are carrying out examining the potential for SBMIs to promote the mental wellbeing of children and adolescents in Rwanda and Ethiopia [25] by helping us to identify what concretely needs to be included in any intervention to obtain the intended outcomes.

## Methods /Design

To achieve our aims and objectives, we will conduct a scoping review to map scientific theories that underpin mindfulness interventions aimed at promoting pupils’ mental wellbeing, drawing on Peters et al. best practice guidelines for reporting items for the development of scoping review [26]. In doing so, we will take account of the advice of Mhairi Campbell and her colleagues based on their experience of doing a review of theories, given that there are no agreed standards for theoretical reviews [27].

A scoping review suits our purpose because we are not primarily concerned with study designs and findings but are more interested in the theories used to justify the use of mindfulness intervention to promote pupils mental wellbeing. For this reason, there is no need to conduct a methodological appraisal of the studies that will be included in our review because we are specifically interested in what they were looking for, what they *expected* to find, and why, rather than *what* they found. However, to ensure the credibility of the theories, we will limit our search to peer review articles and PhDs.

We will use the Preferred Reporting Items for Systematic Reviews and Meta-Analysis extension for Scoping Reviews (PRISMA-ScR) guidance to ensure rigour and facilitate replication (S1 Table: **Preferred Reporting Items for** Systematic reviews and Meta-Analyses extension for Scoping Reviews (PRISMA-ScR) Checklist). The steps involved are (1) identifying the research question; (2) identifying relevant studies; (3) selecting studies for inclusion; (4) charting the studies; and (5) collating, summarising, and reporting the results.

### Step 1: Identifying the research question

The objective of the review is to provide a comprehensive descriptive map and explanation of the theories that have been used to describe the causal pathway between mindfulness practice and mental wellbeing in SBMIs for pupils aged 7-16 years.

1. What theories and frameworks have been identified and described in the literature?
2. How can these theories be classified?

### Step 2: search for evidence

#### Search techniques

The literature search will be in two phases: searching electronic databases and citation tracking (S2 Table: Search Strategy). We will also identify relevant articles from our previous search for documents for a critical realist synthesis of SBMIs. The documents that meet our inclusion criteria will be downloaded for further screening. The search will be restricted to publications in the English language. The search terms and the databases used are based on the advice of an academic librarian.

#### Databases

We will limit our search to scholarly databases and PhD theses to ensure the credibility of the theories. The following databases will be used: MEDLINE, PsychINFO, Web of Science Core Collection, PubMed Central, Scopus and ProQuest Dissertations and Theses.

#### Search strategy and terms

The search terms we will use are: ‘mindfulness’ or ‘mindful’ and ‘wellbeing’ or ‘mental health’ or’ health’ or ‘mental wellbeing’ or ‘happiness’ or ‘happy’ and ‘mechanisms’ or ‘theory’ or ‘theorisation’ or ‘conceptual’ or ‘conceptualisation’ or ‘concept’ or ‘framework’ or ‘mediators’ or ‘moderators’ or ‘process’ or ‘effects’ or ‘scholarship’ and ‘school’ or ‘classroom’ or ‘education’ and ‘pupil’ or ‘child’ or’ adolescent’ or ‘teen’ or ‘student’. Search terms will be adapted for each database. For example, see Table 1 for the search string for MEDLINE and Scopus.

**Table 1:** Search Terms for MEDLINE and Scopus.

|  |  |
| --- | --- |
| **M**<br>**E**<br>**D**<br>**L**<br>**I**<br>**N**<br>**E** | #1 = ((mindfulness or mindful) and (wellbeing or "mental health" or health or "mental wellbeing" or happiness or happy) and (mechanisms or theory or theorisation or conceptual or conceptualisation or concept or framework or mediators or moderators or process or effects or scholarship) and (school or classroom or education) and (pupil or child or adolescent or teen or student)).ab,kf,ti.<br>#2 = limit 1 to english language |
| **S**<br>**c**<br>**o**<br>**p**<br>**u**<br>**s** | TITLE-ABS-KEY ( ( mindfulness OR mindful ) AND ( wellbeing OR "mental health" OR health OR "mental wellbeing" OR happiness OR happy ) AND ( mechanisms OR theory OR theorisation OR conceptual OR conceptualisation OR concept OR framework OR mediators OR moderators OR process OR effects OR scholarship ) AND ( school OR classroom OR education ) AND ( pupil OR child OR adolescent OR teen OR student ) ) AND ( LIMIT-TO ( LANGUAGE , "English" ) ) |

#### Inclusion/exclusion criteria

We will consider all discussion, scholarship or methodology papers that discuss a named generalisable theory or framework for explaining the causal pathway between mindfulness practice and mental wellbeing in SBMIs that are not just targeted at problem pupils. It is anticipated that some studies will discuss their proposed model within the context of a systematic review.

There has been a proliferation of mindfulness interventions for both clinical and non-clinical populations. Therefore, it is crucial to define the essential characteristics of a mindfulness-based programme (MBP) [1,19]. Rebecca Crane and her colleagues have identified the five essential ingredients in MBPs which we will follow in carrying out this scoping review (Table 2).

**Table 2:**
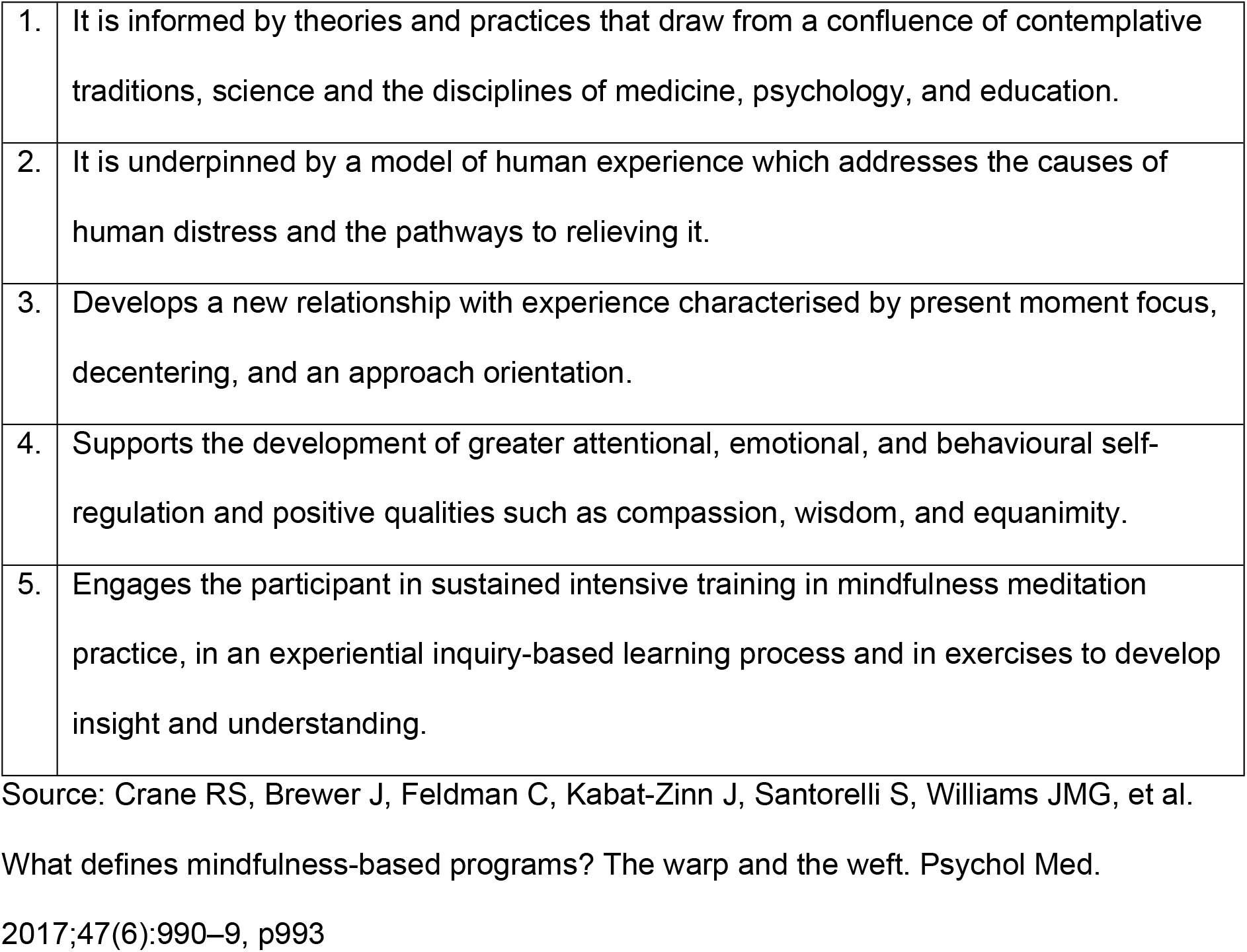
The Five Essential Ingredients of a Mindfulness-based Programme.

#### Inclusion Criteria

- Population: school pupils aged 7-16 years.
- Study design: Theoretical or empirical. The definition of mindfulness used in the study meets the essential requirements (Table 2).
- Documents: from any country or region.
- Any SBMI or theoretical discussion of SBMIs that describes a theory, model or framework connecting mindfulness to mental wellbeing in the design of the intervention or the discussion of the findings.
- Any publication date
- Document type: peer-reviewed theoretical or empirical research papers or PhD thesis
- Language: English.
- The primary or secondary outcome variable (where applicable) measures wellbeing or mental wellbeing.

#### Exclusion Criteria

- There is no reference to a theory, model or framework being useful in the study design or the discussion of outcomes.
- Only discusses theories for understanding how mindfulness interventions work with clinical populations.
- Does not include an outcome measure of wellbeing or mental wellbeing.
- Only includes yoga.
- Mentions mechanisms but does not link them to a theory.
- Not delivered in a school.
- Only included pupils outside the 7-16 year age range.
- Grey literature excluding PhD thesis.
- Not peer-reviewed.
- In languages other than English.

### Step 3: study selection

*Covidence* will be used to manage article screening and data extraction. A four-step search strategy will be utilised.

1. Extracting search results and removing duplicates and citations without abstracts or summaries;
2. A single reviewer will review titles, determining eligibility based on the inclusion and exclusion criteria;
3. Two reviewers will independently review the titles and abstracts remaining, following step two. Differences will be resolved through discussion and, if necessary, by bringing in a third reviewer;
4. All papers left after step 3 will be downloaded and independently evaluated by two reviewers. Differences will be resolved through discussion and, if necessary, by bringing in a third reviewer. Reasons will be recorded for excluding full-text articles and reported in the scoping review.

### Step 4: document appraisal and data extraction

We will extract information from the documents that meet our inclusion criteria into an Excel spreadsheet. The extraction of data will be done under the following headings:

#### General details of the paper

search origin; author and date; title; type of paper (e.g., review paper, primary quantitative study), and type of study.

#### Population focus

(where applicable) country of study, the age range of pupils, and gender.

#### Exposure, mechanism, outcome details

(where relevant) details of the mindfulness intervention.

#### Theories

All theories mentioned in the paper; the name of theories; a detailed description of each theory discussed in the paper; discipline.

If relevant, whether the theory was tested and if findings supported or refuted it; any policy implications explicitly reported.

The extraction tool will be piloted. PA, LD, IS and GN will independently read two documents and complete the extraction table. They will then meet, compare their extraction tables, and agree on necessary modifications.

PA will extract the data from the papers and GN will independently review a 10 per cent sample. The team will meet regularly to discuss the emerging findings and to resolve any differences between PA and GN.

The review to be performed is a focused scoping review to locate and describe existing theoretical models or frameworks; therefore, assessing the methodological quality of the papers is not appropriate.

### Step 5: Analysis and Reporting

Our coding framework will capture the theories linking mindfulness to mental wellbeing. We will identify theory categories, compare theories within each pathway, and compare the theories identified.

Our aim for this scoping review is to present a comprehensive descriptive map and explanation of the theories that have been used to justify the introduction of SBMIS for pupils aged 7-16 years and mental wellbeing. Study characteristics and theories will be presented in a table and summarised using a narrative synthesis approach.

## Data Availability

All the data for the project of which the literature review described in the protocol is part will be made available on completion of the project.

## Ethics

Formal ethics approval is not required for a literature review. However, ethical approval has been obtained from the University of Aberdeen, Addis Ababa University, and the University of Rwanda for the research programme, of which this scoping review forms an integral element.

## Dissemination

We will publish at least one article in a peer review journal reporting the findings from the literature review. Findings from the review will also be disseminated via seminar and conference presentations and podcasts posted on the project website and via social media.

### Authors’ Contributions

PA, GN, and LD contributed to the NIHR grant application. PA led the research design and the protocol’s writing and produced the first draft. GN, IS and LD revised drafts and agreed on the final text of this paper.

## Supporting Information

**S1 Table: Preferred Reporting Items for Systematic reviews and Meta-Analyses extension for Scoping Reviews (PRISMA-ScR) Checklist**

**S2 Table: Search Strategy**

